# Biomechanical Risk Factors for Anterior Cruciate Ligament Injury in Young Female Basketball Players: A pilot Study

**DOI:** 10.1101/2022.07.11.22277460

**Authors:** Akino Aoki, Kohei Koresawa, Yumi No, Masashi Sadakiyo, Satoshi Kubota, Kazuyoshi Gamada

## Abstract

**Objectives:** This study was aimed to reveal the differences in knee valgus angle at landing as a static indicator and wobbling movement of the knee during landing as a dynamic indicator between ACL injury and uninjured athletes.

**Methods:** This study was case-control study. There were 6 female basketball players with ACL injuries and 38 female basketball players without them, whose knee kinematics were measured using 2-dimensional video cameras during single-leg jump landings. The task was performed from 30cm-box. Knee kinematics and wobbling of the knee which was calculated by relative frontal motion to the flexion movement were compared between knees with ACL-injured and uninjured.

**Results:** Six athletes who had confirmed ACL injuries, did not demonstrate significantly different knee valgus angle at initial contact and maximum knee flexion during landing, compared to 38 uninjured athletes. The knee valgus angles at initial contact for injured and uninjured athletes were 12.3° and 14.8° (*p* = 0.15), respectively. Five of six anterior cruciate ligament injured knees presented knee wobbling during landing. Relative frontal motion at 18° knee flexion was significantly greater in athletes with ACL-injured (*p* = 0.02).

**Conclusions:** 84% of ACL injury presented with the knee wobbling and the frontal knee motion was greater with low knee flexion during knee wobbling, while the knee valgus angle was not significantly different. This study suggests that knee wobbling may be a biomechanical and dynamic risk factor for ACL injury in female basketball players.

**Summary Box:** What is already known on this topic – summarise the state of scientific knowledge on this subject before you did your study and why this study needed to be done

Biomechanical risk factor for ACL injury was revealed as dynamic knee valgus and low knee flexion which increases ACL strain. However, previous study focused only static index which are knee angle at initial contact or maximum. This study aimed to establish new dynamic index for screening of ACL injury.

What this study adds – summarise what we now know as a result of this study that we did not know before

Although a previous study identified knee valgus angle and knee valgus moment as predictors of ACL injury, many athletes who demonstrates knee valgus motion does not suffer ACL injury. Cadaveric studies show that ACL strain did not increase when knee valgus occurred with slowed knee flexion movement. We identified an abnormal knee movement involving the dynamic knee valgus with low knee flexion, which we call “knee wobbling.”

How this study might affect research, practice or policy – summarise the implications of this study

ACL injury has been difficult to predict; however, we found that knee wobbling, which is new parameter of abnormal knee movement, including rapid knee valgus/varus, is a potential predictor of ACL injury.

## INTRODUCTION

Anterior cruciate ligament (ACL) injury is a severe sports injury^1^. Annually, 250,000 cases of ACL injury are reported in the United States^2^ alone. The medical cost for one case is approximately $17,000^3^. Nationwide, the cost of reconstructive surgery and rehabilitation for treatment of ACL injuries represents an economic loss of approximately $4.2 billion annually. The incidence of ACL injury is especially higher among female adolescents^4^. Female athletes frequently suffer ACL injuries in noncontact situations that are typically caused by an external load upon the knee joint during a landing, slowing, or pivoting^5,6^. Despite efforts at prevention internationally, the incidence of ACL injury in female athletes has not decreased^7^. There is still no consensus on why the incidence of ACL injury among female athletes is so high, although gender differences, including anatomical, hormonal, and neuromuscular factors are thought to affect the incidence of ACL injury^8^. There is no incontrovertible evidence showing that anatomical or hormonal factors directly or indirectly lead to incidence of ACL injury. Only neuromuscular factors are modifiable as preventative methods, which led researchers to screen athletes with biomechanical testing. Many neuromuscular risk factors associated with ACL injury have been proposed, including an increase in the valgus knee angle^10^, tibial internal rotation^11^, anterior tibial shear force,^12^ hip joint valgus^13^, and adduction^14^ or a decrease in the knee joint flexion angle^12^ or trunk bending angle^15^ during athletics. However, Bahr et al.^9^ claimed that no screening test can predict ACL injury and that no intervention study supports the efficacy of screening. This would be caused since ACL injury occurs with dynamic loads and factors. Establishing dynamic parameters is required for more effective screening methods in order to identify high-risk athletes.

Previous screening methods focused only on an increasing valgus angle in snapshots. The knee valgus angle at IC during a drop vertical jump (DVJ) was reportedly significantly higher in those with an ACL injury than those without^10^. However, many uninjured subjects presented the same amount of knee valgus angle; therefore, there were many false positive subjects in the study^10^. Krosshaug et al.^18^ showed that knee valgus angle during DVJ did not predict ACL injury. These studies analyzed only a snapshot of knee movement during landing, and dynamic knee motion during single leg activity has not been investigated. ACL injury often occurs during deceleration with slight knee flexion (<25°), valgus, and with internal or external rotation^16,17^. From an analysis of knee kinematics at the time of ACL injury in female athletes, using a model-based image matching technique, Koga et al.^16^ showed that the knee valgus angle increased by 12°, while the knee flexion angle increased by only 1° between the initial contact (IC) of the foot during the movement that caused the ACL injury and 40 ms after IC. This study showed that both increasing knee valgus angle and disappearing flexion movement occurred simultaneously at ACL injury. Detecting the combined phenomenon would be necessary for considering risk factors of ACL injury. Only one study examined dynamic knee movement as an index for fluency of knee motion during single leg hops^19^. Roos et al.^19^ showed that knee fluency, meaning without dynamic knee valgus or varus movement, was decreased in ACL-deficient knees more than ACL-intact and ACL-reconstructed knees. Accordingly, combined slight knee flexion and dynamic valgus would be considered a risk factor. However, no study has established the assessment of the dynamic parameter and has identified the association between the new parameter and ACL injury as a potential risk factor.

Better screening methods also need to reproduce the injury mechanism with single-leg tasks. Single-leg activities should be investigated, because many ACL injuries occur during high-impact activity with one leg^6^. However, DVJs with both legs have been utilized in the majority of previous studies. Although some previous studies that utilized single-leg landings, these studies showed only maximum knee valgus or flexion angle^20,21^. This is not adequate to assess dynamic knee control during single-leg activities. In our preliminary study, 24 female athletes performed single-leg jump landings (SLJL). This was an appropriate task for single-leg, high-impact activities. Repeated valgus/varus knee motion (knee wobbling) was observed, and this may be used as an index of dynamic parameters. We speculate that knee wobbling at low knee flexion during SLJL is an unstable dynamic knee movement that may be associated with ACL injury. The objective of this pilot study was to determine differences in knee valgus angle and dynamic knee joint motion during single-leg landings in female basketball players, comparing ACL-injured and uninjured athletes by following them for 3 years. The hypotheses of this study were that (1) there is no difference in the knee valgus angle between ACL-injured and ACL-uninjured knees during single-leg landings, and (2) knee wobbling movement is more frequent in ACL-injured knees than ACL-uninjured knees. We utilized a case-control design to compare knee kinematics between the ACL-injured and uninjured groups by recording kinematic data during SLJL in female basketball athletes at baseline, and by gathering data about non-contact ACL injuries during up to three years of follow-up.

## METHODS

### Subjects

This is a case-control study (Level of Evidence: 3). The study commenced after obtaining approval of the ethics committee of Institutional Review Board of our institution and informed, written consent of all subjects and their parents. Recruitment was conducted from 2010 to 2013. Inclusion criteria were: (1) athletes participating in basketball team sports (competition level); (2) 12-18 years old; (3) female; (4) healthy. Subjects who had any neurological diseases, orthopedic diseases, or communication disorders at the time of data measurement were excluded from the study. Forty-four subjects of two groups agreed to participate in this study. After recruitment and measurement, information on the incidence of non-contact ACL injuries was collected for two or three years. At follow-up, six ACL injuries had occurred. All ACL injuries were diagnosed by an orthopedic surgeon based on magnetic resonance imaging (MRI) findings. ACL injuries that occurred with no contact with another person or object at the time of injury were defined as non-contact ACL injuries. Participants were instructed to report all knee injuries to their coaches and the coaches were instructed to relay the information to the investigators. One investigator continued to contact coaches. Participants played only basketball during the follow-up period.

### Data Acquisition

Subjects wore T-shirts and spats and each subject had six reflective markers attached to her body for video analyses. The reflective markers were 10 mm in diameter and were attached at the greater trochanter, the middle of the patella, and the lateral malleolus on both legs. Subjects performed SLJL with both legs, landing with one leg after a maximum vertical jump from a single-leg standing position on the floor. Two digital video cameras (Casio Computer Co., Ltd., Japan) were used at 30 Hz. Cameras were located in the front and ipsilateral side, 350 cm from the subject and 93 cm from the floor. The knee valgus and flexion angle during SLJL were measured for lower limb kinematics in each frame. The reliability of knee valgus angle for a two-dimensional video analysis at 30 was sufficient to correlate with motion capture during this SLJL^22^.

### Data Processing

We used an Ulead Video Studio 11 (Corel Japan Ltd., Japan) to convert video images into static images at 30 frames/s, and computed the knee valgus and flexion angle in each frame. We then used ImageJ 1.86 (National Institutes of Health, United States) to measure each joint angle on static images. The knee valgus and flexion angle were obtained by deducting the angle formed by a line connecting the greater trochanter, the middle of the patella and the lateral malleolus from 180°. The analyzed phase during SLJL was between IC to maximal knee flexion (MKF). IC was defined as the point when a part of the foot touched the floor, while the MKF was defined as when the knee flexion angle reached its maximum value. All measurements and data analysis were performed by different trained investigators.

Relative frontal motion (RFM) was used as an index of knee valgus/varus movement relative to flexion movement during landing. More precisely, RFM was calculated by the amount of valgus/varus movement in the frontal plane during 1/30 second divided by the amount of flexion movement in the sagittal plane (Figure 1). In order to calculate knee movement at low knee flexion, the knee flexion and valgus angle at IC was set as 0°. Then, the predictive valgus angle at 6°, 12°, 18°, 24° and 30° for standardized knee flexion angle was calculated based on two time points. Positive RFM indicated valgus movement, while negative RFM denoted varus. Positive and negative RFM on successive jumps were interpreted as knee wobbling (Figure 2a). The number of RFM’s between IC and MKF was counted. On the other hand, those with only positive or negative RFM’s were excluded (Figure 2b). The method we developed during a previous study for detecting knee valgus/varus movement with RFM was further validated in the present study^23^.

**Figure 1.**
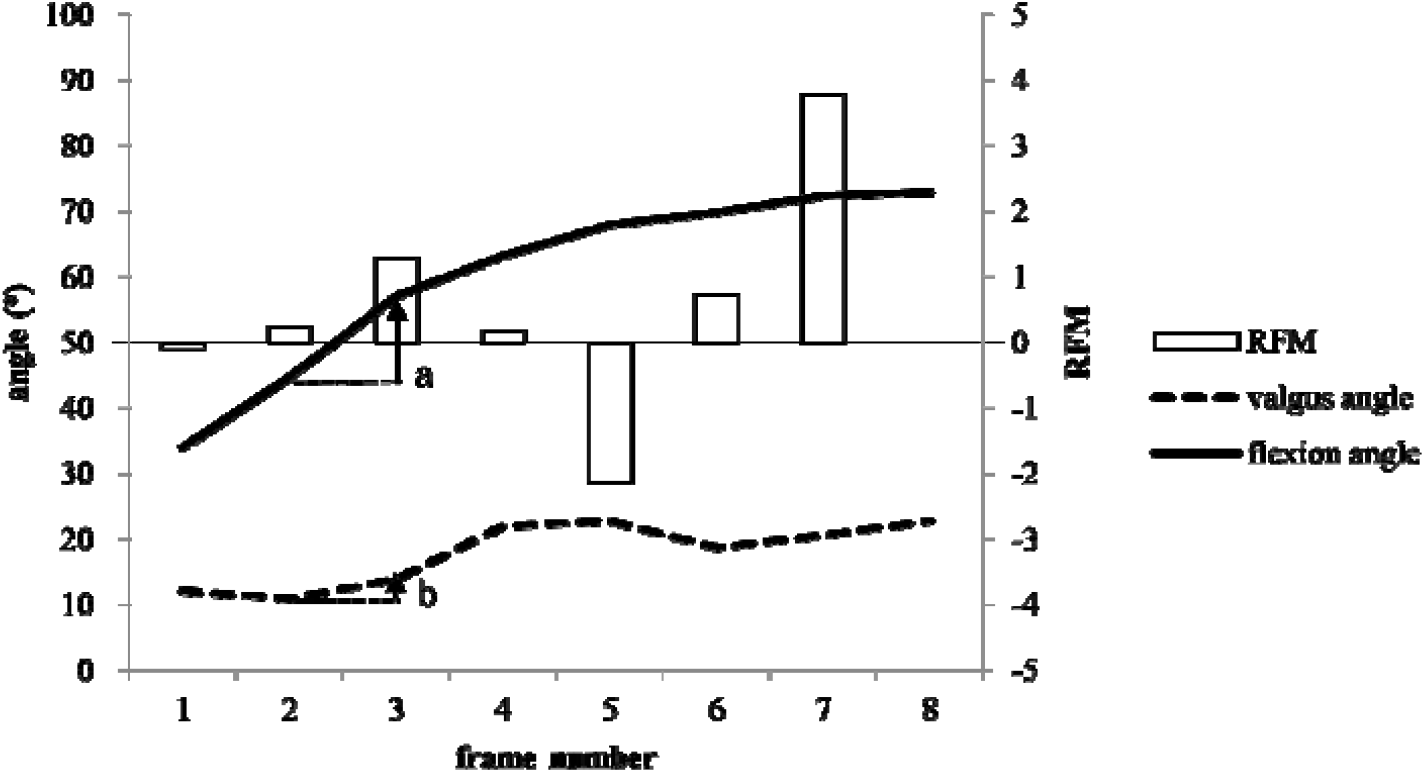
Determination of relative frontal motion (RFM) This graph shows one kinematic dataset from a single-leg landing. RFM was calculated as the amount of valgus/varus movement in the frontal plane (a) divided by the amount of flexion in the sagittal plane (b) during 1/30 second.

**Figure 2.**
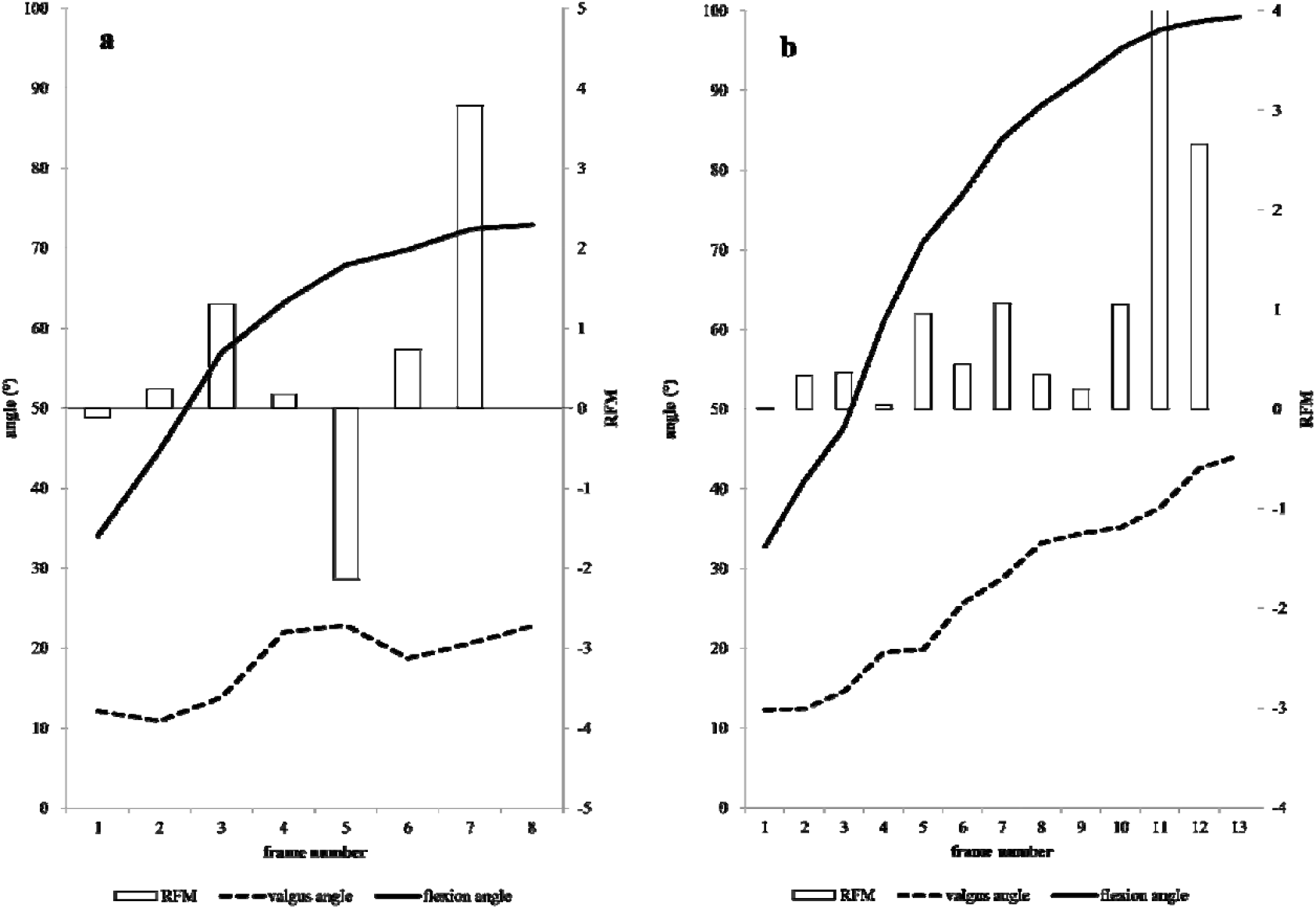
Kinematic data showing repeated knee wobbling and no knee wobbling. (a) Repeated positive and negative RFMs were present. This RFM crossed the zero line between Frames 4 and 5 and between Frames 5 and 6. (b) An example of kinematic data with no knee wobbling. Only positive RFM values are present. This RFM never crossed the zero line.

### Statistical Analyses

Subjects were divided into two groups: ACL-injured and uninjured groups. We computed the mean and standard deviation (SD) of valgus knee angle and flexion angle at IC and MKF and the data presented a normal distribution. T-tests with Bonferroni adjustment were used as parametric tests to compare groups. Chi-square tests were used to assess differences in knee wobbling between ACL-injured and uninjured knees. We used Predictive Analytics Software (PASW) Statistics 18 (Statistical Product and Service Solutions (SPSS), Inc., United States) for statistical analysis, and set the significance level at α < 0.05.

## RESULTS

Six athletes suffered ACL injury after the three-year follow-up period. Table 1 shows the profiles of the six subjects with ACL injuries. There were no significant differences between the six ACL-injured and 38 uninjured subjects in terms of age (ACL-injured: 15.5 ± 1.2 years; uninjured: 16.1 ± 0.3 years; *p* = 0.28), height (ACL-injured: 163.6 ± 4.8 cm; uninjured: 159.5 ± 5.8 cm; *p* = 0.11), weight (ACL-injured: 51.7 ± 6.5 kg; uninjured: 52.7 ± 6.0 kg; *p* = 0.71) (Table 2).

| ACL-injured subject | Age range at time of injury (years) | Injury scene |
| --- | --- | --- |
| Case 1 | 16-18 | Game |
| Case 2 | 16-18 | Practice |
| Case 3 | 16-18 | Game |
| Case 4 | 16-18 | Game |
| Case 5 | 16-18 | Recreation |
| Case 6 | 12-15 | Game |

**Table 2.** Physical characteristics of subjects with and without ACL injuries

|  | ACL-injured subjects | Uninjured subjects | *p* value |
| --- | --- | --- | --- |
| n (persons) | 6 | 38 |  |
| Age (years) <sup>a</sup> | 15.5 ± 1.2 | 16.1 ± 0.3 | 0.28 |
| Height (cm) <sup>a</sup> | 163.6 ± 4.8 | 159.9 ± 5.8 | 0.11 |
| Weight (kg) <sup>a</sup> | 51.7 ± 6.5 | 52.7 ± 6.0 | 0.71 |

<sup>a</sup>: Mean value ± standard deviation

Inter-group comparisons of kinematics during SLJL were performed on the two groups: 6 knees with ACL injuries belonging to 6 subjects and 76 uninjured knees belonging to 38 subjects. Ten uninjured knees were difficult to analyze due to missing markers and were excluded from this study. No differences were detected between the six ACL-injured and 66 uninjured knees in terms of knee valgus, flexion at IC, or MKF (Table 3). Accordingly, no difference in knee joint motion was discernible between the ACL-injured and uninjured knees.

**Table 3.**
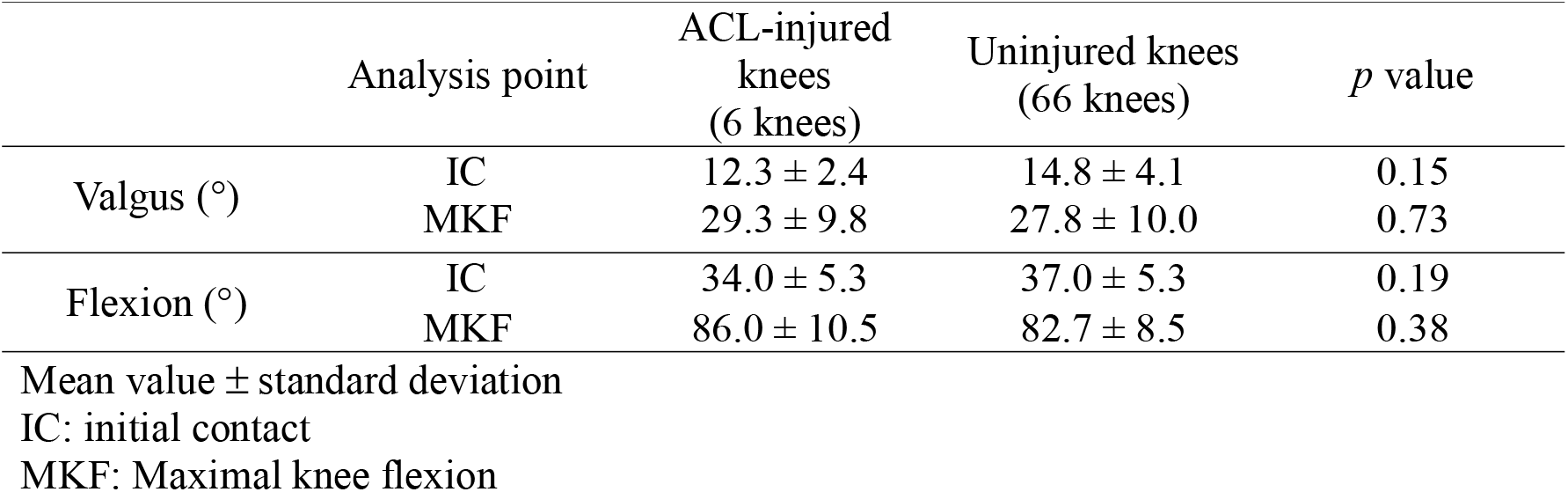
Valgus knee angle and flexion angles at initial contact and maximal knee flexion

Wobbling of the knee was observed in many subjects in both ACL-injured and uninjured knees during SLJL. Although there is no significant difference by chi-square test (*p* = 0.55), the rate of occurrence of knee wobbling was larger in five of the six ACL-injured knees than 41 of the 66 uninjured knees, in at least one jump. No significant difference in relative frontal motion (RFM) was detected between ACL-injured and uninjured knees (Table 4). The RFM’s for ACL-injured knees and uninjured knees at 12°, 18°, 24°, and 30° of knee flexion were 0.09 ± 0.43 and 0 ± 0.25 (*p* = 0.66), 0.42 ± 0.52 and 0.10 ± 0.31 (*p* = 0.02), 0.23 ± 0.37 and 0.13 ± 0.31 (*p* = 0.45), 0.16 ± 0.46 and 0.13 ± 0.48 (*p* = 0.88), respectively. Therefore, the RFM was significantly greater only at 18° in ACL-injured knees.

**Table 4.** RFM at each knee flexion angle

|  | Analysis point<br>(knee flexion<br>angle) | ACL-injured<br>knees<br>(6 knees) | Uninjured<br>knees<br>(66 knees) | *p* value |
| --- | --- | --- | --- | --- |
| RFM<sup>a</sup> | 12° | $0.09 \pm 0.43$ | $0 \pm 0.25$ | 0.66 |
| | 18° | $0.42 \pm 0.52$ | $0.10 \pm 0.31$ | 0.02 |
| | 24° | $0.23 \pm 0.37$ | $0.13 \pm 0.31$ | 0.45 |
| | 30° | $0.16 \pm 0.46$ | $0.13 \pm 0.48$ | 0.88 |

<sup>a</sup>: Mean value ± standard deviation
RFM: relative frontal motion

## DISCUSSION

The objective of this study was to determine the difference in valgus knee angle and knee joint motion between knees that had sustained ACL injuries and those that had not, during SLJL in middle school and high school female basketball players during three years. The knee valgus angle at IC and MKF did not differ significantly between ACL-injured knees and uninjured knees, and abnormal knee joint motion was present in most ACL-injured knees and uninjured knees, but especially in ACL-injured knees.

A combination of low knee flexion and valgus motion was observed in ACL-injured knees^24^. Therefore, screening tasks for ACL injury risks should induce such movement. Valgus angle at landing has been the parameter most evaluated as a potential risk factor^10,16,17^. A previous study indicated that an increase in the knee valgus angle and valgus moment during landing are considered risk factors for non-contact ACL injury^10^. However, many uninjured subjects presented the same knee valgus angle as the injured subjects^10^. In the current study, the knee valgus angle showed no difference between ACL-injured and uninjured knees in either IC or MKF. Thus, we concluded that the knee valgus angle alone during landing is insufficient to portend ACL injury. Another parameter examined was knee flexion angle at IC during landing. The knee flexion angle at the time of ACL injury was between 5 and 25° during single-leg landing or deceleration^25^. The current study found that the knee flexion angle at IC during SLJL was 34.0° for ACL-injured knees and 37.0° for uninjured knees. In a previous study using a motion capture system, the knee flexion angle at IC during a bilateral vertical drop was reportedly 31° ^26^. Therefore, knee flexion was greater during single-leg landing than bilateral-leg landing. Despite the high flexion angle at landing during SLJL, landing movement with one leg would be more appropriate as a screening task to detect risk factors than that with both legs during a landing task considering that ACL injury occurs during single-leg activities.

RFM may serve as an index of dynamic knee motion in the frontal and sagittal planes. Previous studies screened knee kinematics using only snapshots at IC or MKF^10^. The RFM used in this study was the amount of change in the frontal plane angle during 1/30 s divided by the amount of change in the sagittal plane angle. The greater RFM at 18° in ACL-injured knees suggests a sudden valgus or varus and slow flexion movement. Therefore, RFM clearly demonstrated the quality of dynamic valgus/varus movement during landing when compared with snapshots of knee angle at IC or MKF. Decreasing knee flexion movement during landing should be taken into consideration to detect high-risk athletes. A cadaveric study showed that the combined knee valgus external torque at slight knee flexion angle induced excessive ACL strain. In the current study, five of the six ACL-injured knees showed knee wobbling, which led us to consider this as a risk factor for ACL injury. The cause of this abnormal knee movement is unknown, and the mechanism of knee wobbling and the association between wobbling and ACL injury should be studied in the future. Since the flexion angle exceed 30° during the SLJL, knee wobbling over a smaller flexion range should be induced and evaluated in future studies.

This study had several limitations. First, we used video cameras for kinematic analyses. Nagano et al.^27^ performed a study to obtain correlations based on regression analysis between two-dimensional and three-dimensional knee kinematics and there was a moderate association between the two methods. Since two-dimensional evaluation is a simple, low-cost technique, we believe that it is a more useful method than three-dimensional analysis when conducting a large-scale study of risk factors. Also, RFM was calculated based on the amount of change and it is not dependent on the method of capture. Second, conclusions of this study cannot be generally applied to males or to athletes involved in other sports. However, this study did follow up female young basketball athletes for a long time and this is a strong point. Third, although the sample size was not large, the post-hoc power was medium to high, which were 0.41 for t-tests and 0.50 for chi-square tests. In the future, there needs to be a larger study with high accuracy, including athletes involved in other high-risk sports, for example, soccer or football.

## Conclusion

No differences in the knee valgus or flexion angles at IC or MKF during the SLJL was detected between the ACL-injured and uninjured knees. Knee wobbling may be more common in ACL-injured athletes and greater RFM at 18° during landing may be a potential risk factor for ACL injury. Future studies should evaluate knee kinematics in a smaller knee flexion range during movement using a different screening task.

## Supporting information

title page

Acknowledgment

author disclosure

STROBE checklist

## Data Availability

All data produced in the present work are contained in the manuscript

