## Supplementary material for "Biomechanical Risk Factors for Anterior Cruciate Ligament Injury in Young Female Basketball Players: A pilot Study": title page

Meaningful Titles:

Biomechanical Risk Factors for Anterior Cruciate Ligament Injury in a Young Female Basketball Player

Running Head:

Risk Factors for ACL Injury

Authors:

Akino Aoki, PT, PhD^1^

Kohei Koresawa, PT, MSc^2^

Yumi No, PT^3^

Masashi Sadakiyo, PT^4^

Satoshi Kubota, PT, PhD^5^

Kazuyoshi Gamada, PT, PhD^6^

^1^ Department of Physical Therapy, International University of Health and Welfare, Chiba, Japan

^2^Department of Rehabilitation, Sanjo Sports Medicine and Orthopaedic Surgery Clinic, Kagawa, Japan

^3^Department of Rehabilitation, Imamura Orthopaedics, Nagasaki, Japan

^4^Department of Rehabilitation, Sadamatsu Hospital, Nagasaki, Japan

^5^Department of Physical Therapy, School of Health Sciences, Tokyo International University, Saitama, Japan

^6^Graduate school of Medical Technology and Health Welfare Science, Hiroshima International University, Hiroshima, Japan

Author Contributions Statement:

Akino Aoki contributed to the interpretation data and drafting this paper. Kohei Koresawa and Satoshi Kubota contributed to the research design and the analysis of data. Yumi No and Masashi Sadakiyo contributed to the acquisition of data. Kazuyoshi Gamada contributed to revising this paper and approval of the submitted and final versions.

The study protocol was approved by the Ethics Committee of Sadamatsu Hospital in Nagasaki, Japan. The authors certify that they have no affiliations with or financial involvement in any organization or entity with a direct financial interest in the subject matter or materials discussed in the article.

Corresponding author:

Akino Aoki

4-3 Kozunomori, Narita city, Chiba, Japan

International University of Health and Welfare

Abstract:

Objectives: This study was aimed to reveal the differences in knee valgus angle at landing as a static indicator and wobbling movement of the knee during landing as a dynamic indicator between ACL injury and uninjured athletes.

Methods: This study was case-control study. There were 6 female basketball players with ACL injuries and 38 female basketball players without them, whose knee kinematics were measured using 2-dimensional video cameras during single-leg jump landings. The task was performed from 30cm-box. Knee kinematics and wobbling of the knee which was calculated by relative frontal motion to the flexion movement were compared between knees with ACL-injured and uninjured.

Results: Six athletes who had confirmed ACL injuries, did not demonstrate significantly different knee valgus angle at initial contact and maximum knee flexion during landing, compared to 38 uninjured athletes. The knee valgus angles at initial contact for injured and uninjured athletes were 12.3° and 14.8° (*p* = 0.15), respectively. Five of six anterior cruciate ligament injured knees presented knee wobbling during landing. Relative frontal motion at 18° knee flexion was significantly greater in athletes with ACL-injured (*p* = 0.02).

Conclusions: 84% of ACL injury presented with the knee wobbling and the frontal knee motion was greater with low knee flexion during knee wobbling, while the knee valgus angle was not significantly different. This study suggests that knee wobbling may be a biomechanical and dynamic risk factor for ACL injury in female basketball players.
