## Supplementary material for "Biomechanical Risk Factors for Anterior Cruciate Ligament Injury in Young Female Basketball Players: A pilot Study": author disclosure

**Author Disclosure Statement:**

In the normal course of the conduct and publication of medical research, associations commonly exist that might prevent an author from reporting research with complete objectivity, or that might be perceived as doing so. The AJSM has adopted a policy of "transparency" in such matters, so that any circumstance that might be perceived as interfering with objectivity is openly declared. Individuals should note that a "potential conflict of interest" describes a circumstance or association, not a behavior, and does not imply that one's behavior has actually been influenced by the potentially conflicting circumstance.

Each author should answer the following 5 questions directly related to the submitted work:

1. Have you received any funding or sponsorship for this study, including funds or materials (e.g., surgical supplies, specimens)?

None.

2. Have you, in the past five years, received the following from an organization that may in any way gain or lose financially from the results of your study:

- Reimbursement for attending a symposium?
- A fee for speaking or for organizing an educational program?
- Payment to write or review a paper?
- Funds for research?
- Funds for one of your employees?
- Royalties or fees for consulting?

None.

3. Have you, in the past five years, been employed by or on the Board of an organization that may in any way gain or lose financially from the results of your study?

None.

4. Do you hold any stocks or bonds in an organization that may in any way gain or lose financially from the results of your study? (This excludes stocks or bonds that may be held in mutual funds.)

None.

5. Do you, or any entity with which you are affiliated, currently hold any patents that are relevant to the study in any way, or are you currently applying for or planning to apply for such a patent?

None.

Each author should answer the following 4 questions regarding disclosures not directly related to the current work:

6. Do you have any other potential financial conflict of interest?

None.

7. Do you have any affiliation with any commercial entity or organization not related to this particular study, but related to orthopaedic surgery or another branch of medicine?

None.

8. Do you have any non-reimbursed relationships that may be seen as a conflict of interest?

None.

9. Does your spouse, parent, child, or sibling have a relationship as described above in questions 1 through 8?

None.

**Any payments to authors over $500 appearing in the Open Payments Database (https://openpaymentsdata.cms.gov) for the last 5 years should be included on this form. When disclosing potential conflicts, you do not need to provide dollar amounts, but please be sure to indicate the company and nature of payment: educational support, royalties, hospitality payments (food and lodging), etc.**

If none of the authors have answered, "Yes," to any of the above 9 questions, please mark option number 1 below, "No Potential Conflicts of Interest declared."

If one or more authors have answered, "Yes," to any of these 9 questions, please mark option number 2 below.

☑1. Please insert "I (we) declare that we have no conflicts of interest in the authorship or publication of this contribution."

Or

__ 2. Please insert "One or more of the authors has declared a potential conflict of interest as specified in the AJSM Conflict of Interest statement."

Please specify below

- Which of the potential conflicts are being declared
- The specific author(s) declaring a potential conflict of interest
- Any other circumstance, which is not covered in the potential conflicts of interest listed above, that you feel might be perceived as constituting a potential conflict of interest and wish to voluntarily declare

____________________________________________________

____________________________________________________

**Statement of Sponsorship**

Please describe the sponsorship of your study, including any direct or indirect financial support and any donations of equipment, services, supplies, specimens, use of testing facilities, etc.

**Statement of Authorship**

All authors have contributed substantially to the conception, design, analysis, and/or interpretation of the data in this manuscript and will take public responsibility for the content.

__A.A__ Initials of corresponding author

Has anyone not listed as an author on this paper contributed to the conduct or analysis of this study or the editing, writing, or translation of this manuscript?

____ Yes

☑No

If yes, please identify the individual(s) or entity and describe their role. Also, please include disclosure for these individuals.

**Description of Publication Restrictions**

Have all authors had unrestricted access to all the data of this study?

_____Yes

☑ No

If No, please describe the restrictions.
Only personal data is restricted to one of the co-authors.

Have the named authors had the final authority over the content of this paper?

☑Yes

____No

If No, please identify the party who had the final authority.

**Statement of Related Publications**

Manuscripts must not have been published elsewhere in any language or substantially overlap or duplicate published work or work that is currently in preparation. Manuscripts must not be under simultaneous consideration by any other publication, before or during the peer-review process. Manuscripts should cite any other work by one or more of the co-authors that is relevant to the subject matter of the current submission or that used any of the same subjects, animals, or specimens being reported in the current submission. This includes manuscripts that are currently under preparation, are being considered by journals, are accepted for publication, or already published. In any of these cases, the relationship to the current submission should be made clear.

Do any authors have a related or similar publication in preparation, in press, or published?

______ Yes

☑ No

If yes, has this paper been referenced in the work submitted for consideration?

_____ Yes

______ No

If No, please cite or describe these works and explain the reasons for not citing them.

AJSM does not allow for publication of manuscripts already uploaded to preprint servers, and this would therefore disqualify the material from further consideration. Has this paper been uploaded to a preprint server?

______ Yes

☑ No

Signature of Corresponding Author (on behalf of all authors) ________________________

Title of paper: _ Biomechanical Risk Factors for Anterior Cruciate Ligament Injury in Young Female Basketball Players: A pilot Study____

MS Number: _81476207717___

Date: _7 Jul 2022_
