## Supplementary material for "Biomechanical Risk Factors for Anterior Cruciate Ligament Injury in Young Female Basketball Players: A pilot Study": STROBE checklist

STROBE Statement—checklist of items that should be included in reports of observational studies

| | Item No. | Recommendation | Page No. | Relevant text from manuscript |
|---|---|---|---|---|
| **Title and abstract** | 1 | (*a*) Indicate the study's design with a commonly used term in the title or the abstract | 1 | case-control study |
| | | (*b*) Provide in the abstract an informative and balanced summary of what was done and what was found | 1 | knee wobbling as an ACL injury risk factor |
| **Introduction** | | | | |
| Background/rationale | 2 | Explain the scientific background and rationale for the investigation being reported | 3 | combined slight knee flexion and dynamic valgus would be considered a risk factor. |
| Objectives | 3 | State specific objectives, including any prespecified hypotheses | 4 | to determine differences in knee valgus angle and dynamic knee joint motion during single leg landing in female basketball players |
| **Methods** | | | | |
| Study design | 4 | Present key elements of study design early in the paper | 5 | case-control study |
| Setting | 5 | Describe the setting, locations, and relevant dates, including periods of recruitment, exposure, follow-up, and data collection | 5-10 | |
| Participants | 6 | (*a*) *Cohort study*—Give the eligibility criteria, and the sources and methods of selection of participants. Describe methods of follow-up<br>*Case-control study*—Give the eligibility criteria, and the sources and methods of case ascertainment and control selection. Give the rationale for the choice of cases and controls<br>*Cross-sectional study*—Give the eligibility criteria, and the sources and methods of selection of participants | 5 | At follow-up, six ACL injury was occurred. |
| | | (*b*) *Cohort study*—For matched studies, give matching criteria and number of exposed and unexposed<br>*Case-control study*—For matched studies, give matching criteria and the number of controls per case | 5 | 298 athletes with non-ACL injury was excluded due to no video data of single leg jump landing and 44 athletes was included as non-ACL injury group. |
| Variables | 7 | Clearly define all outcomes, exposures, predictors, potential confounders, and effect modifiers. Give diagnostic criteria, if applicable | 7 | Relative frontal motion (RFM) was used as an index of knee valgus/varus movement relative to the flexion movement |
| Data sources/ measurement | 8* | For each variable of interest, give sources of data and details of methods of assessment (measurement). Describe comparability of assessment methods if there is more than one group | 7 | The subjects performed SLJL with both legs, which is the landing with one leg after a maximum vertical jump. |
| Bias | 9 | Describe any efforts to address potential sources of bias | 7 | All data analysis was performed by a trained investigator who was another one by examiner of measurement |
| Study size | 10 | Explain how the study size was arrived at | 18 | the post-hoc power was 0.41 and 0.50 for t-test and chi-square test, respectively |

Continued on next page

| | | | | |
|---|---|---|---|---|
| Quantitative variables | 11 | Explain how quantitative variables were handled in the analyses. If applicable, describe which groupings were chosen and why | 7 | RFM was calculated by an amount of valgus/varus movement in the frontal plane during 1/30 second divided by the amount of flexion movement in the sagittal plane |
| Statistical methods | 12 | (*a*) Describe all statistical methods, including those used to control for confounding | | Not applicable |
| | | (*b*) Describe any methods used to examine subgroups and interactions | | Not applicable |
| | | (*c*) Explain how missing data were addressed | 11 | no video data of single leg jump landing |
| | | (*d*) *Cohort study*—If applicable, explain how loss to follow-up was addressed<br>*Case-control study*—If applicable, explain how matching of cases and controls was addressed<br>*Cross-sectional study*—If applicable, describe analytical methods taking account of sampling strategy | | Not applicable |
| | | (*e*) Describe any sensitivity analyses | | Not applicable |
| **Results** | | | | |
| Participants | 13* | (a) Report numbers of individuals at each stage of study—eg numbers potentially eligible, examined for eligibility, confirmed eligible, included in the study, completing follow-up, and analysed | 5,11 | Six suffered ACL injury during the three-year follow-up period.<br>Ten uninjured knees were difficult to analyze and were excluded from this study. |
| | | (b) Give reasons for non-participation at each stage | 5 | no video data of single leg jump landing |
| | | (c) Consider use of a flow diagram | 6 | flow diagram shows |
| Descriptive data | 14* | (a) Give characteristics of study participants (eg demographic, clinical, social) and information on exposures and potential confounders | 11 | There were no significant differences between the six ACL-injured and 38 uninjured subjects in demographic data. |
| | | (b) Indicate number of participants with missing data for each variable of interest | 5 | no video data of single leg jump landing in 282 |
| | | (c) *Cohort study*—Summarise follow-up time (eg, average and total amount) | | Not applicable |
| Outcome data | 15* | *Cohort study*—Report numbers of outcome events or summary measures over time | | Not applicable |
| | | *Case-control study*—Report numbers in each exposure category, or summary measures of exposure | 12 | 6 knees belonging to 6 ACL-injured subjects and 76 knees belonging to 38 uninjured subjects |
| | | *Cross-sectional study*—Report numbers of outcome events or summary measures | | Not applicable |
| Main results | 16 | (*a*) Give unadjusted estimates and, if applicable, confounder-adjusted estimates and their precision (eg, 95% confidence interval). Make clear which confounders were adjusted for and why they were included | 12 | mean and SD |
| | | (*b*) Report category boundaries when continuous variables were categorized | | Not applicable |
| | | (*c*) If relevant, consider translating estimates of relative risk into absolute risk for a meaningful time period | | Not applicable |

Continued on next page

| | | | | |
|---|---|---|---|---|
| Other analyses | 17 | Report other analyses done—eg analyses of subgroups and interactions, and sensitivity analyses | | not applicable |
| **Discussion** | | | | |
| Key results | 18 | Summarise key results with reference to study objectives | 17 | abnormal knee joint motion was present in most of the ACL-injured knees and uninjured knees, especially in ACL-injured knees. |
| Limitations | 19 | Discuss limitations of the study, taking into account sources of potential bias or imprecision. Discuss both direction and magnitude of any potential bias | 18 | This study had several limitations. |
| Interpretation | 20 | Give a cautious overall interpretation of results considering objectives, limitations, multiplicity of analyses, results from similar studies, and other relevant evidence | 18 | Knee wobbling may be present in ACL-injured athletes with higher prevalence. |
| Generalisability | 21 | Discuss the generalisability (external validity) of the study results | 18 | the subjects in this study cannot be generally applied to males or athletes involved in other sports. |
| **Other information** | | | | |
| Funding | 22 | Give the source of funding and the role of the funders for the present study and, if applicable, for the original study on which the present article is based | | not applicable |

*Give information separately for cases and controls in case-control studies and, if applicable, for exposed and unexposed groups in cohort and cross-sectional studies.

**Note:** An Explanation and Elaboration article discusses each checklist item and gives methodological background and published examples of transparent reporting. The STROBE checklist is best used in conjunction with this article (freely available on the Web sites of PLoS Medicine at http://www.plosmedicine.org/, Annals of Internal Medicine at http://www.annals.org/, and Epidemiology at http://www.epidem.com/). Information on the STROBE Initiative is available at www.strobe-statement.org.
